# IC3 Protocol – A novel digital health method for monitoring cognition after stroke

**DOI:** 10.1101/2023.08.23.23294469

**Authors:** Dragos C. Gruia, William Trender, Peter Hellyer, Soma Banerjee, Joseph Kwan, Henrik Zetterberg, Adam Hampshire, Fatemeh Geranmayeh

## Abstract

**Introduction:** Stroke is a major cause of death and disability worldwide, frequently resulting in persistent cognitive deficits among survivors. These deficits negatively impact recovery and therapy engagement, and their treatment is consistently rated as high priority by stakeholders and clinicians. Although clinical guidelines endorse cognitive screening for post-stroke management, there is currently no gold standard approach for identifying cognitive deficits after stroke, and clinical stroke services lack the capacity for long-term cognitive monitoring and care. Currently available assessment tools are either not stroke-specific, not in-depth or lack scalability, leading to heterogeneity in patient assessments.

**Methods and Analysis:** To address these challenges, a cost-effective, scalable, and comprehensive screening tool is needed to provide a stroke-specific assessment of cognition. The current study presents such a novel digital tool, the Imperial Comprehensive Cognitive Assessment in Cerebrovascular Disease (IC3), designed to detect both domain-general and domain-specific cognitive deficits in patients after stroke with minimal input from a health professional. To ensure its reliability, we will utilise multiple validation approaches, and aim to recruit a large normative sample of age-, gender-, and education-matched UK-based controls. Moreover, the IC3 assessment will be integrated within a larger prospective observational longitudinal clinical trial, where post-stroke cognition will be examined in tandem with brain imaging and blood biomarkers to identify novel multimodal biomarkers of recovery after stroke. This study will enable deeper cognitive phenotyping of patients at a large scale, whilst identifying those with highest risk of progressive cognitive decline, as well as those with greatest potential for recovery.

**Ethics and dissemination:** This study has been approval by South West - Frenchay Research Ethics Committee (IRAS 299333), and authorized by the UK’s Health Research Authority. Study registration. The study is registered as an observational trial under NCT05885295.

## Strengths and limitations

- This study addresses post stroke cognitive deficits, a major clinical problem.
- Accurately predicting the trajectory of cognitive recovery is a largely unmet clinical and scientific need.
- We developed a novel digital screening tool called the Imperial Comprehensive Cognitive Assessment in Cerebrovascular Disease (IC3), that is not only cost-effective and scalable, but can also provide deep phenotyping of stroke-specific cognitive deficits with minimal input from a health professional.
- The IC3 tool will be used within a longitudinal observational study for identifying novel brain imaging and blood biomarkers of recovery after stroke.
- This study will enable large-scale cognitive profiling and monitoring of patients, improving our ability to identify those at the highest risk of progressive cognitive decline and those with the greatest potential for recovery.

## Introduction and Rationale

Stroke is a leading cause of death and disability globally, with frequent cognitive sequalae affecting three-quarters of survivors [1]. The spectrum of cognitive deficits includes background ‘vascular cognitive impairment’ and more domain-specific deficits, such as aphasia which affects a third of patients [2]. Collectively, these impairments negatively impact post-stroke recovery and engagement with therapy [3]. Furthermore, there is a high failure rate of post-stroke cognitive rehabilitation trials, due to our poor understanding of individual patient-level mechanisms of post-stroke cognitive recovery and the ensuing inadequate stratification of both treatment responders and non-responders in the same clinical trials. It is therefore important to understand the factors that affect cognitive recovery post-stroke, a topic increasingly recognised by funders and stakeholders as a high priority research area [4, 5].

Although clinical guidelines have been recently updated to recommend cognitive screening for post-stroke management, there is currently no gold standard approach for identifying cognitive deficits after stroke [6]. Clinicians and researchers often select cognitive tests depending on preference, familiarity and availability, leaving little prospect for generalizability of their findings. Moreover, clinical stroke services do not have the capacity to provide long-term cognitive monitoring and care for stroke survivors who have unpredictable recovery trajectories [3, 7]. This challenge is further compounded by the lack of cost-effective cognitive tools suitable for long-term monitoring, and the reliance of such monitoring on resource-stretched healthcare professionals.

Availability of a cost-effective, reliable, and comprehensive screening tool that provides a stroke-specific deep phenotyping of cognition would be a game-changer. In the clinical setting, it would improve the detection rate and monitoring of cognitive deficits post stroke, and in the research setting it would facilitate much needed large-scale population-based mechanistic studies aimed at understanding the mechanisms of cognitive recovery. Here, we present such novel digital adaptive tool: The Imperial Comprehensive Cognitive Assessment in Cerebrovascular Disease (IC3). IC3 is a digital assessment designed to require minimal input from a clinician in detecting both domain-general and domain-specific cognitive deficits in patients after stroke. To ensure its reliability, several validation studies will be conducted against established clinical screening tools, and normative samples will be computed using representative age-, gender- and education-matched UK-based controls.

In line with recent efforts in identifying biomarkers of post-stroke recovery [8], the IC3 tool will be integrated within a larger prospective longitudinal clinical study, where post-stroke cognition will be examined in tandem with brain imaging and novel blood biomarkers. This study is entitled “Understanding factors affecting cognitive function in cerebrovascular disease” and will hereafter be referred to as the ‘*main study*’.

## Methods and Design

### Aims

1. Develop an adaptive, scalable, self-administered, digital comprehensive cognitive screening tool to detect cognitive impairments in patients with cerebrovascular disease. The IC3 will:

1. Detect both domain-general (e.g. executive function) and domain-specific (e.g. aphasia, apraxia, neglect) deficits post-stroke [9].
2. Allow monitoring to occur at scale, in a cost-efficient manner, as test administration can occur independent of trained professionals.
3. Minimise the effects of neglect or aphasia on cognitive assessments.
4. Have high test-re-test reliability for repeated-testing longitudinally.
2. Validate the IC3 against commonly used cognitive screening tools.
3. Use IC3 in a cohort of stroke survivors to map the cognitive recovery trajectory over the course of the first year after stroke.
4. Identify novel biomarkers of cognitive outcome (at 1 year) and recovery trajectory through the main study using:

1. Demographics, and comorbid physical, neuropsychological and socioeconomic factors
2. Structural and functional MRI brain imaging, to assess metrics of cerebrovascular disease load, stroke lesion topology and brain network dynamics.
3. Blood biomarkers of Alzheimer’s disease (phosphorylated tau forms and amyloid D42/40 ratio), neuroaxonal injury (neurofilament light, NFL and brain-derived tau), and astrocytic activation (glial fibrillary acidic protein, GFAP). The study will characterise the time course of these biomarkers over the first year after stroke and relate these to MRI measures of axonal injury and brain atrophy, as well as cognitive and functional outcomes.

### IC3 Assessment Design and Development

Cognitive tests. The IC3 assessment covers 22 short tasks, spanning a wide range of cognitive domains (Table 1) followed by several clinically-validated questionnaires, designed to be completed in under 60-70 minutes. IC3 has in-built automated break reminders every 20 minutes with the additional option to take an unscheduled break at any time. IC3 is available via a web-browser on any modern device (smartphone, tablet, computer/laptop) by clicking on a link created by the study team. The IC3 test is implemented via Cognitron, a state-of-the art platform for remote neuropsychological testing rapidly being adopted by large scale population studies both in the UK and internationally [10].

**Table 1:** Tasks tested in the IC3 and their associated cognitive domain.

| Cognitive Domain |  |  | Task Name |
| --- | --- | --- | --- |
| Primary | Secondary | Tertiary |  |
| Attention | Spatial ability | Space | Pear Cancellation (Visual-spatial ability) |
|  |  | Object |  |
|  | Auditory sustained attention |  | Auditory Attention Task |
|  | Visual sustained attention | Simple upper-limb movement | Simple Reaction Time |
| Executive | Task switching and attention |  | Non-Verbal Trail Making (Task Switching) |
|  | Reasoning | Mental shifting and flexibility | Rule Finding |
|  |  |  | Odd One Out (Pattern Recognition) |
|  |  | Spatial planning | Blocks |
| Language | Speech production | Phonology and semantics | Naming |
|  |  | Phonology | Repetition |
|  |  | Phonology, fluency, and semantics | Spoken Picture Description |
|  |  | Reading | Reading |
|  | Semantics | Verbal semantics | Semantic Judgment |
|  | Comprehension (word-picture or word-sentence matching) | Auditory and Orthographic processing | Language comprehension |
| Memory | Long Term |  | Orientation |
|  | Short Term | Verbal working memory | Digit span |
|  |  | Visuo-spatial working memory | Spatial span |
|  |  | Visuo-spatial memory | Paired Associates Learning |
|  | Episodic | Visuo-spatial | Task Recall |
|  |  | Verbal | Target Word Recall from Auditory Attention Test |
| Number Skills |  |  | Graded Calculation |
| Praxis | Ideomotor action semantics |  | Gesture Recognition |
| Motor ability | Left-right upper-limb movement | Attention | Choice Reaction Time (click on left or right side of the screen) |
|  | Multi-directional upper-limb movement |  | Motor Control (click on moving target) |

The digital nature of the IC3 affords scalability in cognitive monitoring by being usable in both the clinical setting and home environment in the absence of a trained clinician. As well as reducing healthcare costs, this feature promotes accessibility of the assessment for physically disabled patients in whom attendance to healthcare or research setting is difficult. Compared to pen-and-paper tests, the IC3 test administration is standardised, more detailed response metrics per individual tests are provided (e.g. accuracy, reaction time and trial-by-trial variability), and real-time patient scores are calculated automatically against a large age-, education- and sex-matched control sample. The IC3 specifications are summarised in Figure 1.

**Figure 1.**
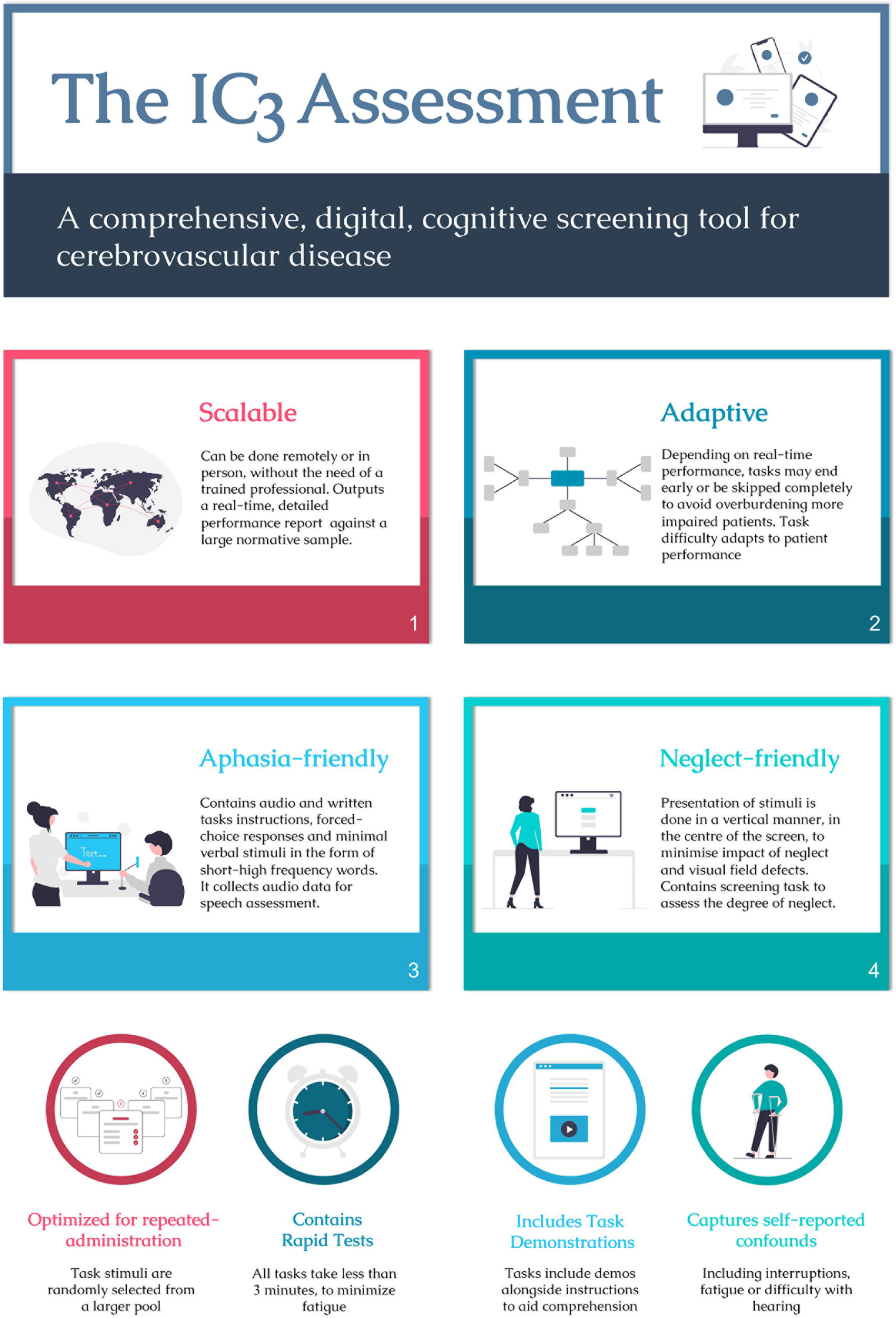

Demographic and neuropsychiatric questionnaires. A number of health questionnaires and modified versions of relevant clinically-validated questionnaires are completed by the patient or carers, including Apathy Evaluation Scale [6], Fatigue Severity Scale [11], Geriatric Depression Scale [12], Instrumental Activities of Daily Living (See Table 2). Upon completion of the cognitive tests, the participants also fill in modified versions of relevant clinically-validated questionnaires, including Modified short version of Apathy Evaluation Scale [6], Modified short version of Fatigue Severity Scale [11], Short form of the Geriatric Depression Scale [12], Instrumental Activities of Daily Living [13].

**Table 2:** Questionnaire items included in the IC3.

|  | **Range/Categories** |
| --- | --- |
| Age | 0-100 |
| Biological sex | Male/Female |
| Years of education | Forced choice answers scaled to 0-21 |
| Handedness | Likert scale ranging from 1 to 5, where 1 is right-handed, 3 is ambidextrous and 5 is left-handed |
| Hand used for making responses | Left/Right |
| Hand impairments | Yes/No |
| English proficiency | Native, Excellent but non-native, moderate level, low-level |
| Ethnicity | White, Asian, Black, Arab, Mixed, Other |
| Occupational status | Unemployed, long-term sick, retired, student, self-employed, employed |
| Annual income | £0 - £100,000+ |
| Existence of stroke risk factors | Diabetes, high-blood pressure, high cholesterol, heart disease, kidney disease, alcohol dependency, overweight, smoker, ex-smoker |
| Existence of Medical Conditions-<br>(followed by more probing questions if an option is selected) | Stroke, other neurological problems, depression, anxiety |
| ***If participant indicates a history of stroke*** |  |
| How long ago was the stroke? | Date/Time |
| How many strokes | 0-10 |
| Type of stroke | Ischemic, Haemorrhagic, TIA, Subarachnoid Haemorrhage, I don't know |
| Problems caused by the stroke | Language, Memory, Concentration, Movement of the right arm/leg, Movement of the left arm/leg, Vision, other |
| Movement | Difficulties standing, sitting, and walking |

At the end of the assessment the participant will be given a graphical, easy-to-understand diagram that highlights their performance against normative data (Figure 2). The normative data will be based on single time-point data collected remotely from 5,000 healthy controls. For tasks with normally distributed data, mild, moderate and severe impairment will be classed using suggested cut-offs of −1.5, −2.0 and −2.5 SD below the mean respectively. For tasks with ceiling effects, 90^th^, 95^th^ and 99^th^ percentiles are the suggested cut-offs.

**Figure 2.**
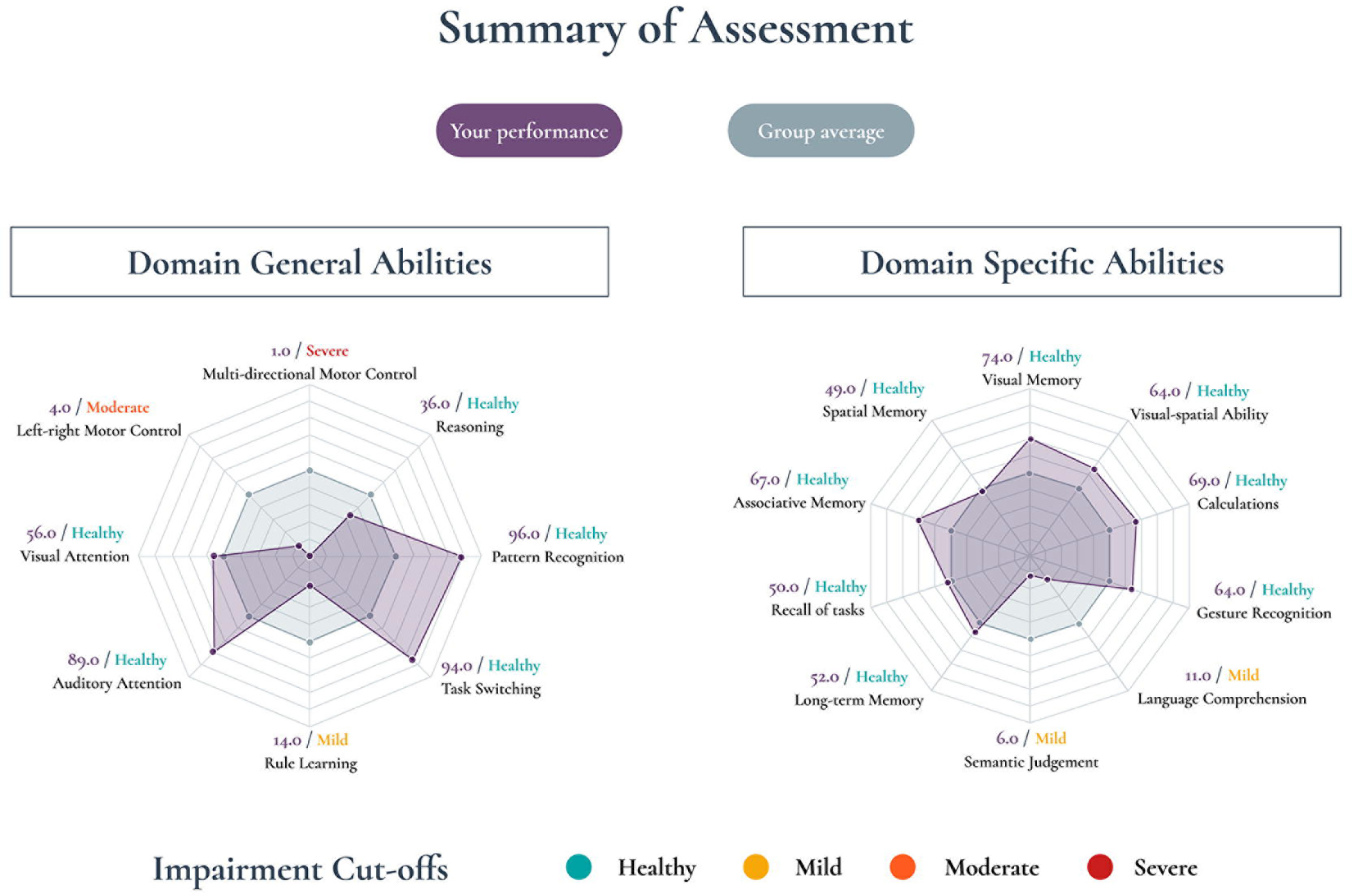

### Main IC3 study design

This prospective observational longitudinal study aims to recruit 300 patients with acute stroke and obtain repeated measures of cognition and psychosocial status at baseline, 3-months, 6-months and 12-months post ictus. IC3 is currently a single-site study at Imperial College Healthcare NHS Trust, England, and commenced recruitment in February 2022. In addition to the cognitive monitoring, all patients will be invited to take part in MRI brain imaging and blood biomarker sub-studies (Figure 3; see sub-studies for more details).

**Figure 3.**
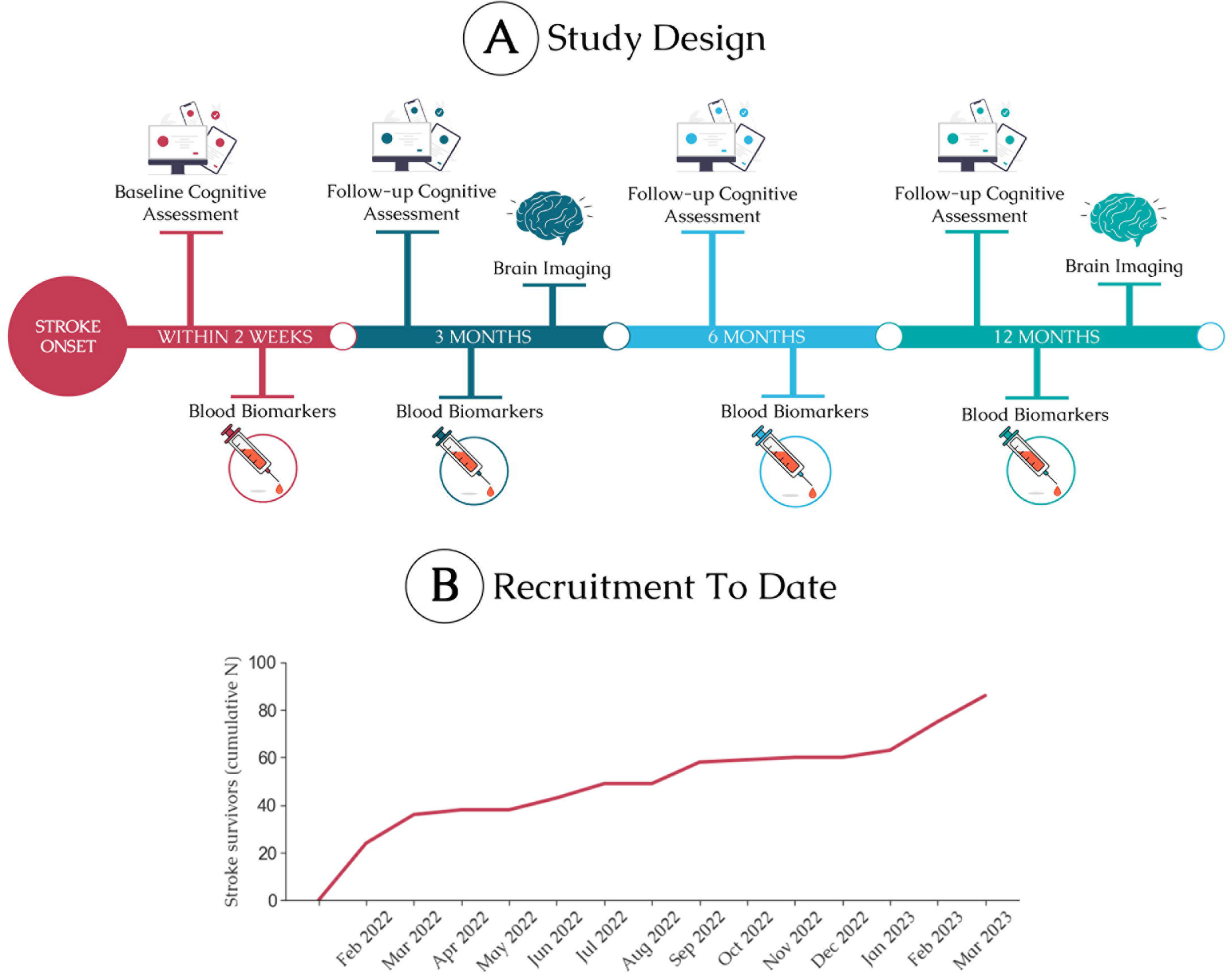

IC3 validation sub-study. A minimum of 100 patients will undergo clinically validated pen and paper cognitive screens used routinely in clinical practice including MOCA [14], star cancellation task [15] and/or the OCS. Sensitivity, specificity analyses, as well as convergence and divergence analyses will be performed. Test-retest reliability analyses will estimate the impact of learning effects on performance, and compare remote administration with in-person delivery of the assessment.

MRI sub-study. Clinically acquired brain imaging at the time of the acute stroke (including FLAIR and DWI sequences) will be obtained. All 300 patients will be invited to undergo additional brain MRI imaging at 3 and 12 months using a Siemens Verio 3T scanner at the Imperial College Clinical Imaging Facility. MRI measures will be used as prognostic indicators for cognitive trajectory. MRI data will be collected from additional 50 age-matched control participants for comparison. The following sequences will be acquired at each session over 60-90 minutes (breaks included):

1. Structural MRI: T1 (1 mm^3^ resolution for volumetric analyses of lesion volume and brain atrophy); Susceptibility Weighted Imaging (0.8×0.8×3.0 mm^3^ for identification of microbleeds); T2 FLAIR (1 mm^3^ for detecting white matter hyperintensity volume (WMHV)); and NODDI (an advanced multi-shell diffusion MRI with 90 directions and 9 b_0_ images) to assess axonal integrity and tissue microstructure. Lesions will be defined using established standards [16].
2. Functional MRI: Resting-state and breath-hold paradigm will measure dynamic regional brain activity and vascular reactivity respectively [17].

Blood biomarkers sub-study. Blood samples will be obtained at baseline (0-15 days post-stroke), and at 3, 6 and 12 months from 200 patients. Two EDTA (plasma) and two SST (serum) samples will be collected. Sample processing involves centrifugation at 2000g for 10 minutes at 4°C, followed by secure storage at −80°C. Additional single PAXgene sample will be collected at one time point for DNA storage for future analysis of apolipoprotein E (APOE) genotype and polygenic risk score measures. Patient identity is kept confidential. The primary plasma biomarker of interest will be NFL [18]. Additional markers including GFAP (astrocytic activation [19]), brain-derived tau (a novel marker for neuronal injury [20]), phosphorylated tau forms and AD42/40 ratio (both biomarkers for Alzheimer’s pathologies[21]) will be quantified. Testing will be performed at University College London via a Quanterix Simoa analyser to provide ultrasensitive measurement of concentrations.

### Patient and public involvement statement

The IC3 assessment was developed and refined during a 6-months iterative process of design modifications based on patient and user feedback. We conducted feedback sessions with stroke survivors part-way through the development of our assessment to ensure that (1) our cognitive assessments are intuitive, (2) the instructions are comprehensible, (3) patients can complete the assessment with minimum supervision, and (4) the burden of assessments are acceptable. Based on these sessions, we made several improvements to the design of the assessment, primarily focusing on improving the user interface, user experience and the instructions. We also involved multiple age-matched members of the public, without a diagnosis of stroke, to ensure that our design changes were effective and to identify any issues with performing the assessment remotely. Finally, we disseminated the study design through local public outreach events (e.g., educational events for older adults, health advocacy charities) and incorporated the received feedback in the end product.

### Study Population

Participant identification, recruitment. Supplementary Table 1 outlines the patient inclusion and exclusion criteria. Patients will be recruited from the Imperial College Healthcare NHS Trust by study personnel and the clinical team. Our goal is to recruit patients as early after stroke as is practical, and ideally within fifteen days to facilitate acute blood biomarker assessment. Control participants will be aged > 18 and without history of neurological or severe psychiatric illness.

### Outcome measures

The primary outcome measure for cognition will be the IC3-derived accuracy measures for each primary cognitive domain listed in Table 1 at 1 year post ictus. Secondary outcome measures will be (a) MOCA score [14], (b) functional measures including NIH Stroke Scale [22], Modified Rankin Scale [23], Instrumental Activities of Daily Living [13] and (c) psychological measures via the Geriatric Depression Scale [12] and Apathy Evaluation Scale [6].

### Data analysis plan

Cognition. Dimensionality reduction techniques, such as Principal Component Analysis, graph-based methods and hierarchical clustering will examine the inter-relationship between cognitive domains and will be used to derive latent variables. These will be used to relate to blood and imaging biomarkers. Such cognitive profiling will also form the basis for future real-time optimization of the IC3 task presentation based on patients’ performance.

To assess the trajectories of functional, clinical, and psychological outcome measures, we will employ linear mixed effect models. These models will include cognitive measures as the main predictors, together with standard covariates such as age, sex, lesion, and initial cognitive deficit.

Blood biomarkers. NFL, GFAP, amyloid, and tau biomarkers will be analysed to describe their dynamics after acute stroke and to compare differences between groups. Additionally, longitudinal changes in these biomarkers will be assessed within patients using non-parametric tests, where appropriate. The relationship between blood biomarker levels and continuous outcome measures at 1 year (e.g., cognitive scores), will be investigated using mixed-effects linear regression. For binarized outcome measures (e.g., favourable or unfavourable MRS scores), logistic regression will be employed.

Brain imaging. The following analyses will be performed to derive imaging predictors of cognition and relate to blood biomarker levels: i) WMHV evaluated using FSL BIANCA tool (https://fsl.fmrib.ox.ac.uk/fsl/fslwiki/BIANCA); ii) stroke lesion topology manually delineated on FLAIR images; iii) NODDI measures of axonal injury and tissue microstructure (FA, neural density and orientation dispersion index) analysed in NODDI MATLAB Toolbox; iv) Cortical thickness measured using Freesurfer; v) vascular reactivity analysis [17]; vi) Resting-state fMRI analyses of the effect of stroke on functional brain networks. These measures will be compared between groups. Longitudinally, each patient will act as their own ‘control’ in linear regressions accounting for age, sex, initial stroke severity, lesion volume, and time. Finally, structural equation modelling will be used to determine the relationship between demographics, comorbidities, blood and brain imaging biomarkers and cognitive and functional outcome measures.

### Sample Size

The primary blood biomarker predictor is NFL. Based on preliminary reported group differences [18], we expect 140 patients at baseline, and 60 patients at 3-months post stroke (when peak NFL is expected to occur), will be able to detect group differences in NFL levels between patients and controls (Wilcoxon Mann Whitney test, Cohens D=0.38 and 0.58 respectively, alpha=0.05, power=0.8, 1-tailed, allocation ratio 2:1). A much smaller sample size of 56 will be enough to detect correlations between NFL and time of sampling of the acute blood: (r =0.10 [18], alpha=0.05, power=0.8, 1-tailed).

The primary imaging outcome measure is WMHV. A sample size of 80 will be sufficiently powered to detect correlation between NFL and WMHV based on preliminary findings [24] (r=0.32, alpha 0.05, power 0.95). A sample size of N=70 will be sufficiently powered to detect relationship between peak plasma NFL levels and binary functional outcomes (based on similar effects observed after acute brain injury (R = 0.28 [25], a=0.05, power=0.9). Expecting a 20-25% loss to follow-up rate (due to patient factors and technical factors), the stated sample size of 200 in blood biomarker sub-study is sufficiently powered to detect all the above-mentioned effects.

A secondary blood biomarker predictor is raised phosphorylated tau. Based on the mean and standard deviations reported for longitudinal change in phosphorylated tau (N=374) in population of elderly cognitively normal population [26], a sample size of N=65, will have 95% power in detecting any true change in mean over 1-year period (a=0.05, effect size 0.41). Thus, the stated sample size is sufficiently powered to detect longitudinal change in phosphorylated tau.

## Discussion

Cognitive impairments are highly prevalent after stroke, having detrimental implications to recovery [1]. Identifying, monitoring and managing these deficits is consistently voted the highest priority by stakeholders [5]. Given the heterogeneity of stroke and its co-morbid conditions (such as aging, neurodegeneration and background small vessel disease), there is a need for large-scale longitudinal studies that enable accurate predictions of recovery and pave the way for personalised therapy allocation. Previous longitudinal studies of post-stroke outcomes have failed to identify reliable biomarkers of recovery [7]. On the one hand, this is due to a narrow focus on only limited contributors to recovery (e.g. lesion topology), despite the fact that stroke recovery is known to be inherently multi-modal [8]. On the other hand, this is due to using brief, shallow cognitive assessments in small-sized samples. Our main longitudinal study addresses this gap by coupling emerging blood and brain imaging biomarkers with a novel scalable digital technology (IC3) that allows in-depth remote monitoring of cognition, thus providing a multi-modal account of recovery.

Moreover, current stand-alone cognitive screening tools commonly used in routine clinical care are either not stroke-specific, or not comprehensive. Commonly used tools, such as MOCA [14], MMSE [27], and ACE-R [28] are tailored to detect deficits in neurodegenerative dementias and are not suitable for patients with stroke, who often have domain-specific as well as domain-general deficits [9]. Additionally, increasingly used cognitive screens designed specifically for stroke, such as the OCS, OCS Plus and CASP are not in-depth enough to allow a deep cognitive phenotyping and likely to miss the milder end of the severity spectrum. Assessments that are both stroke-specific and comprehensive, such as BCOS, are too resource-intensive for routine clinical practice or longitudinal research studies into post-stroke cognition. The IC3 assessment tool developed and applied in this study addresses these shortcomings, building the foundation for routine detailed monitoring of cognition and for scalable large-scale population-based studies of post-stroke cognitive impairment.

The blood biomarker sub-studies are expected to yield novel findings concerning the role of emerging biomarkers in neurodegenerative dementias, specifically amyloid and tau entities, as well as the role of biomarkers of neuroaxonal injury, such as NFL and GFAP, in determining cognitive outcomes post-stroke. The study will investigate the longitudinal dynamics of these biomarkers in the post-acute stroke period. The integration of both brain imaging biomarkers (e.g., white matter integrity, tissue microstructure, vascular reactivity, small vessel disease load) and blood biomarkers is expected to result in a cutting-edge and easily applicable predictive models for post-stroke cognition. The IC3 study is expected to make a significant contribution to a growing number of well-designed biomarker studies aimed at post-stroke cognition (such as R4VAD [29] and Discovery [30]) which have the potential to significantly advance the treatment of a disease that affects millions of people globally every year. These studies will enable more precise targeting of cognitive rehabilitation to individuals who are most at risk of progressive decline and those who have the highest potential for recovery.

## Contributions

FG designed the study and obtained funding. DCG, FG, AH, HZ, WT, PH, SB and JK contributed to the data collection plan. FG, DCG, AH, WT and PH developed IC3 platform. DCG wrote the first draft of the manuscript. All authors reviewed and approved the manuscript.

## Data management and regulatory approval

All data collected will be pseudo-anonymised and securely stored on university server. Blood samples will be stored at the Imperial College Clinical Research Facility and Imperial College Healthcare NHS Trust. Data management procedures will be compliant with both Imperial College London guidelines and GDPR regulations.

## Funding

This research is funded by MRC MR/T001402/1. Infrastructure support was provided by the NIHR Imperial Biomedical Research Centre and the NIHR Imperial Clinical Research Facility. The views expressed are those of the author(s) and not necessarily those of the NHS, the NIHR or the Department of Health and Social Care.

### Informed Consent and Ethical Approval

This study has been approval by South West - Frenchay Research Ethics Committee (IRAS 299333), and authorized by the UK’s Health Research Authority. All participants satisfying the inclusion and exclusion criteria will be approached and consented. If a patient is unable to provide fully informed consent, for example due to severe aphasia, we will assent with permission from a consultee. Patients that have been assented will be reconsented if they are subsequently able to provide full informed consent. The consent procedure will be carried out in strict compliance with national legislation and General Data Protection Regulation.

## Data Availability

As the current work only discusses the study's protocol, there is no data available. The authors, however, are happy to share more information about the IC3 software upon reasonable request.

## References

[1] Douiri A, Rudd AG, Wolfe CD. Prevalence of poststroke cognitive impairment: South London stroke register 1995–2010. Stroke 2013; 44: 138–145.

[2] Grönberg A, Henriksson I, Stenman M, et al. Incidence of Aphasia in Ischemic Stroke. Neuroepidemiology 2022; 56: 174–182.

[3] Obaid M, Flach C, Marshall I, et al. Long-Term Outcomes in Stroke Patients with Cognitive Impairment: A Population-Based Study. Geriatrics 2020; 5: 32.

[4] Hill G, Regan S, Francis R, et al. Research priorities to improve stroke outcomes. Lancet Neurol 2022; 21: 312–313.

[5] Pollock A, St George B, Fenton M, et al. Top 10 Research Priorities Relating to Life after Stroke – Consensus from Stroke Survivors, Caregivers, and Health Professionals. Int J Stroke 2014; 9: 313–320.

[6] Quinn TJ, Elliott E, Langhorne P. Cognitive and Mood Assessment Tools for Use in Stroke. Stroke 2018; 49: 483–490.

[7] Boyd LA, Hayward KS, Ward NS, et al. Biomarkers of stroke recovery: Consensus-based core recommendations from the Stroke Recovery and Rehabilitation Roundtable. Int J Stroke 2017; 12: 480–493.

[8] Simpkins AN, Janowski M, Oz HS, et al. Biomarker Application for Precision Medicine in Stroke. Transl Stroke Res 2020; 11: 615–627.

[9] Demeyere N, Riddoch MJ, Slavkova ED, et al. Domain-specific versus generalized cognitive screening in acute stroke. J Neurol 2016; 263: 306–315.

[10] Hampshire A, Hellyer PJ, Soreq E, et al. Associations between dimensions of behaviour, personality traits, and mental-health during the COVID-19 pandemic in the United Kingdom. Nat Commun 2021; 12: 4111.

[11] Krupp LB, LaRocca NG, Muir-Nash J, et al. The fatigue severity scale: application to patients with multiple sclerosis and systemic lupus erythematosus. Arch Neurol 1989; 46: 1121–1123.

[12] Herrmann N, Mittmann N, Silver IL, et al. A validation study of The Geriatric Depression Scale short form. Int J Geriatr Psychiatry 1996; 11: 457–460.

[13] Lawton MP, Brody EM. Assessment of older people: self-maintaining and instrumental activities of daily living. The gerontologist 1969; 9: 179–186.

[14] Nasreddine ZS, Phillips NA, Bédirian V, et al. The Montreal Cognitive Assessment, MoCA: A Brief Screening Tool For Mild Cognitive Impairment. J Am Geriatr Soc 2005; 53 695–699.

[15] Friedman PJ. The Star Cancellation Test in acute stroke. Clin Rehabil 1992; 6: 23–30.

[16] Wardlaw JM, Smith EE, Biessels GJ, et al. Neuroimaging standards for research into small vessel disease and its contribution to ageing and neurodegeneration. Lancet Neurol 2013; 12: 822–838.

[17] Geranmayeh F, Wise RJS, Leech R, et al. Measuring vascular reactivity with breath-holds after stroke: A method to aid interpretation of group-level BOLD signal changes in longitudinal fMRI studies. Hum Brain Mapp 2015; 36: 1755–1771.

[18] Pedersen A, Stanne TM, Nilsson S, et al. Circulating neurofilament light in ischemic stroke: temporal profile and outcome prediction. J Neurol 2019; 266: 2796–2806.

[19] Chatterjee P, Doré V, Pedrini S, et al. Plasma Glial Fibrillary Acidic Protein Is Associated with 18F-SMBT-1 PET: Two Putative Astrocyte Reactivity Biomarkers for Alzheimer’s Disease. J Alzheimers Dis JAD 2023; 92: 615–628.

[20] Gonzalez-Ortiz F, Turton M, Kac PR, et al. Brain-derived tau: a novel blood-based biomarker for Alzheimer’s disease-type neurodegeneration. Brain J Neurol 2023; 146: 1152–1165.

[21] Zetterberg H, Bendlin BB. Biomarkers for Alzheimer’s disease-preparing for a new era of disease-modifying therapies. Mol Psychiatry 2021; 26: 296–308.

[22] Wityk RJ, Pessin MS, Kaplan RF, et al. Serial assessment of acute stroke using the NIH Stroke Scale. Stroke 1994; 25: 362–365.

[23] van Swieten JC, Koudstaal PJ, Visser MC, et al. Interobserver agreement for the assessment of handicap in stroke patients. Stroke 1988; 19: 604–607.

[24] Gattringer T, Pinter D, Enzinger C, et al. Serum neurofilament light is sensitive to active cerebral small vessel disease. Neurology 2017; 89: 2108–2114.

[25] Graham NSN, Zimmerman KA, Moro F, et al. Axonal marker neurofilament light predicts long-term outcomes and progressive neurodegeneration after traumatic brain injury. Sci Transl Med 2021; 13: eabg9922.

[26] Moscoso A, Grothe MJ, Ashton NJ, et al. Longitudinal associations of blood phosphorylated Tau181 and neurofilament light chain with neurodegeneration in Alzheimer disease. JAMA Neurol 2021; 78: 396–406.

[27] Kurlowicz L, Wallace M. The mini-mental state examination (MMSE). Journal of gerontological nursing 1999; 25: 8–9.

[28] Mioshi E, Dawson K, Mitchell J, et al. The Addenbrooke’s Cognitive Examination Revised (ACE-R): a brief cognitive test battery for dementia screening. Int J Geriatr Psychiatry 2006; 21: 1078–1085.

[29] Wardlaw JM, Doubal F, Brown R, et al. Rates, risks and routes to reduce vascular dementia (R4vad), a UK-wide multicentre prospective observational cohort study of cognition after stroke: Protocol. Eur Stroke J 2021; 6: 89–101.

[30] Rost NS, Meschia JF, Gottesman R, et al. Cognitive Impairment and Dementia After Stroke: Design and Rationale for the DISCOVERY Study. Stroke; 52. Epub ahead of print August 2021. DOI: 10.1161/STROKEAHA.120.031611.

